# Down-regulation of *MALAT1* is a hallmark of tissue and peripheral proliferative T cells in COVID-19

**DOI:** 10.1101/2023.01.06.23284229

**Authors:** Shoumit Dey, Helen Ashwin, Luke Milross, Bethany Hunter, Joaquim Majo, Andrew J Filby, Andrew J Fisher, Paul M. Kaye, Dimitris Lagos

## Abstract

T cells play key protective but also pathogenic roles in COVID-19. We studied expression of long non-coding RNAs (lncRNAs) in COVID-19 T cell transcriptomes by integrating previously published single-cell RNA sequencing datasets. The long intergenic non-coding RNA *MALAT1* was the most highly transcribed lncRNA in T cells, with Th1 cells demonstrating the lowest and CD8+ resident memory cells the highest *MALAT1* expression, amongst CD4+ and CD8+ T cells populations, respectively. We then identified gene signatures that covaried with *MALAT1* in single T cells. A significantly higher number of transcripts correlated negatively with *MALAT1* than those that correlated. Enriched functional annotations of the *MALAT1*-anti-correlating gene signature included processes associated with T cell activation such as cell division, oxidative phosphorylation and response to cytokine. The *MALAT1* anti-correlating gene signature shared by both CD4+ and CD8+ T cells marked dividing T cells in both lung and blood of COVID-19 patients. Focussing on the tissue, we used an independent patient cohort of post-mortem COVID-19 lung samples and demonstrated that *MALAT1* suppression was indeed a marker of MKI67+ proliferating CD8+ T cells. Our results reveal *MALAT1* suppression and its associated gene signature are a hallmark of human proliferating T cells.

## Introduction

T cell plasticity and balance is crucial for protection and pathology (Noack & Miossec, 2014; Pennock et al., 2013). Such opposing roles have been appreciated in various provocations of the immune system and more recently in the progression of COVID-19 (Z. Chen & John Wherry, 2020; Moss, 2022). While antigen-specific T cells may confer protection to SARS-CoV-2 virus (Sekine et al., 2020; A. T. Tan et al., 2021; Z. Wang et al., 2021), lymphopenia is associated with severe COVID-19 (G. Chen et al., 2020; Diao et al., 2020; L. Tan et al., 2020) and exhausted, senescent T cells and those expressing MKI67 (Rupp et al., 2021; Schub et al., 2020), a key proliferation marker, contribute to pathology (Adamo, Chevrier, et al., 2021; Z. Chen & Wherry, 2020; Mazzoni et al., 2021; Shrotri et al., 2021). In CD8+ T cells, a strongly proliferative phenotype correlates with contraction and disappearance of clones (Adamo, Michler, et al., 2021).

Long non-coding RNAs (lncRNAs) are regulatory non-coding RNAs, longer than 200nt. In most cases, lncRNAs show low-medium expression with poor conservation across species often acting as scaffolds for recruitment, sequesters for chromatin-modifiers or RNA binding proteins to specific genomic sites (Engreitz et al., 2016; Guo & Guttman, 2022). LncRNAs may be *cis-* or *trans-*acting wherein the former influences transcription by affecting the loci near their transcription site (enhancer-like) while the latter transcripts leave the transcription site to affect gene expression (mRNA-like) via transcriptional or post-transcriptional mechanisms (Gil & Ulitsky, 2019).

LncRNAs play essential roles in adaptive immunity, particularly in lymphocyte activation, signalling and effector functions (Zeni & Mraz, 2021). For example, lncRNAs such as *lncHSC-2* commit HSCs to lymphoid specification as B or T cells (Luo et al., 2015). T cell development is regulated by Notch1 signalling (Wilson et al., 2001) whose expression is in turn regulated by the lncRNA *NALT1* (Y. Wang et al., 2015). As T cells mature, their activation is triggered by T cell receptors (TCRs) upon MHC mediated antigen presentation that is further modulated by co-stimulatory or co-inhibitory ligands. This activation leads to a switch to glycolysis (W van der Windt Erika L Pearce et al., 2012) which is in turn is influenced by the lncRNA *PVT1* (Fu et al., 2020; Johnsson & Morris, 2014). Indeed, sets of lncRNAs specifically regulate lineage-specific gene expression in activated T cells (Plasek & Valadkhan, 2021) such as Th1 (Petermann et al., 2019), Th2 (Gibbons et al., 2018), Th17 (Braga-Neto et al., 2020) and Treg (Jiang et al., 2017) programmes.

Profiling lncRNA expression in immune cells during response to infection can provide insights into key transcriptional and post-transcriptional mechanisms operating in health and disease. Of note, even though the transcriptomes of tissue and peripheral T cells during responses to infection, and more specifically SARS-CoV-2 have been extensively studied (Stephenson et al., 2021), the study of T cell lncRNA profiles has been limited (Plowman & Lagos, 2021; Yang et al., 2021).

We explored T cell lncRNA profiles from three publicly available datasets from individuals with COVID-19 identifying several lncRNAs that are detectable in lung T cells during infection. We particularly focused on *MALAT1*, a long intergenic non-coding RNA (lincRNA, a sub-class of lncRNAs) remarkably conserved in vertebrates (Gutschner et al., 2013). LincRNAs such as MALAT1 do not overlap with protein coding genes and can have various regulatory effects on gene expression (Ransohoff et al., 2017). Localised in nuclear speckles (Tripathi et al., 2010), *MALAT1* is known to work in a variety of ways, such as through binding splicing factors (Tripathi et al., 2010), controlling function of proteins involved transcription (West et al., 2014), miRNA sequestration (YiRen et al., 2017), and associating with proteins (Sun & Ma, 2019). *MALAT1* has been associated with positively regulating cell cycle progression in cancer tissues (Tripathi et al., 2013) a loss of which impairs cell proliferation (Wang et al., 2016).

*MALAT1* has been shown to regulate T cell function, predominantly in animal models of infection or immunopathology (Kanbar et al., 2022; Liang & Tang, 2020; Masoumi et al., 2019; Xue et al., 2022). In a previous study, in CD4+ T cells, we reported that MALAT1 downregulation is a hallmark of naïve CD4+ T cell activation and that *MALAT1*-/-CD4+ T cells express lower levels of IL-10, an anti-inflammatory cytokine resulting in enhanced inflammation or immunity in experimental models of leishmaniasis and malaria (Hewitson et al., 2020).

Here, we examined COVID-19 single cell RNA sequencing (scRNA seq) datasets from bronchoalveolar lavage (BAL) (Liao et al., 2020; Wauters, Mol, et al., 2021), explant/post-mortem lung cells (Bharat et al., 2020) and peripheral blood (Lee et al., 2020) and discovered that *MALAT1* was negatively correlated with cell cycle progression and proliferation in CD4+ and CD8+ T cells of severe COVID-19 patients. Performing RNAscope on COVID-19 post-mortem lung tissue from individuals who died of COVID-19, we confirmed that MKI67-expressing CD8+ T cells had lower levels of *MALAT1* mRNA *in situ*. Overall, our findings reveal that MALAT1 expression in T cells from COVID-19 patients is linked to a specific gene signature and that low *MALAT1* expression is a hallmark of proliferative T cells.

## Results

### MALAT1 is differentially expressed in CD4+ and CD8+ sub-populations

We integrated T cells BAL scRNAseq datasets (Liao et al., 2020; Wauters, Van Mol, et al., 2021) to look at highly expressed lncRNAs in T cells from healthy volunteers and individuals with COVID-19 (**Methods; Figure 1A**). We found *MALAT1* to be the highest expressed lncRNA with similar distribution in both datasets which is ubiquitously found across all T cells (**Figure 1A**). We then normalised and integrated the two datasets (see **Methods**) and clustered them at a low resolution to infer coarse grained T cell heterogeneity (**Figure 1B, S1**). Cells visualised on UMAP showed both the datasets to be similarly spread across UMAP space indicating similar composition (**Figure 1C left**). We then used cell type metadata (Liao et al., 2020; Wauters, Van Mol, et al., 2021) to obtain a finer grained T cell phenotyping (**Figure 1C middle**). We found that there was marked differences between T cells based on disease severity (**Figure 1C right**).

**Figure 1:**
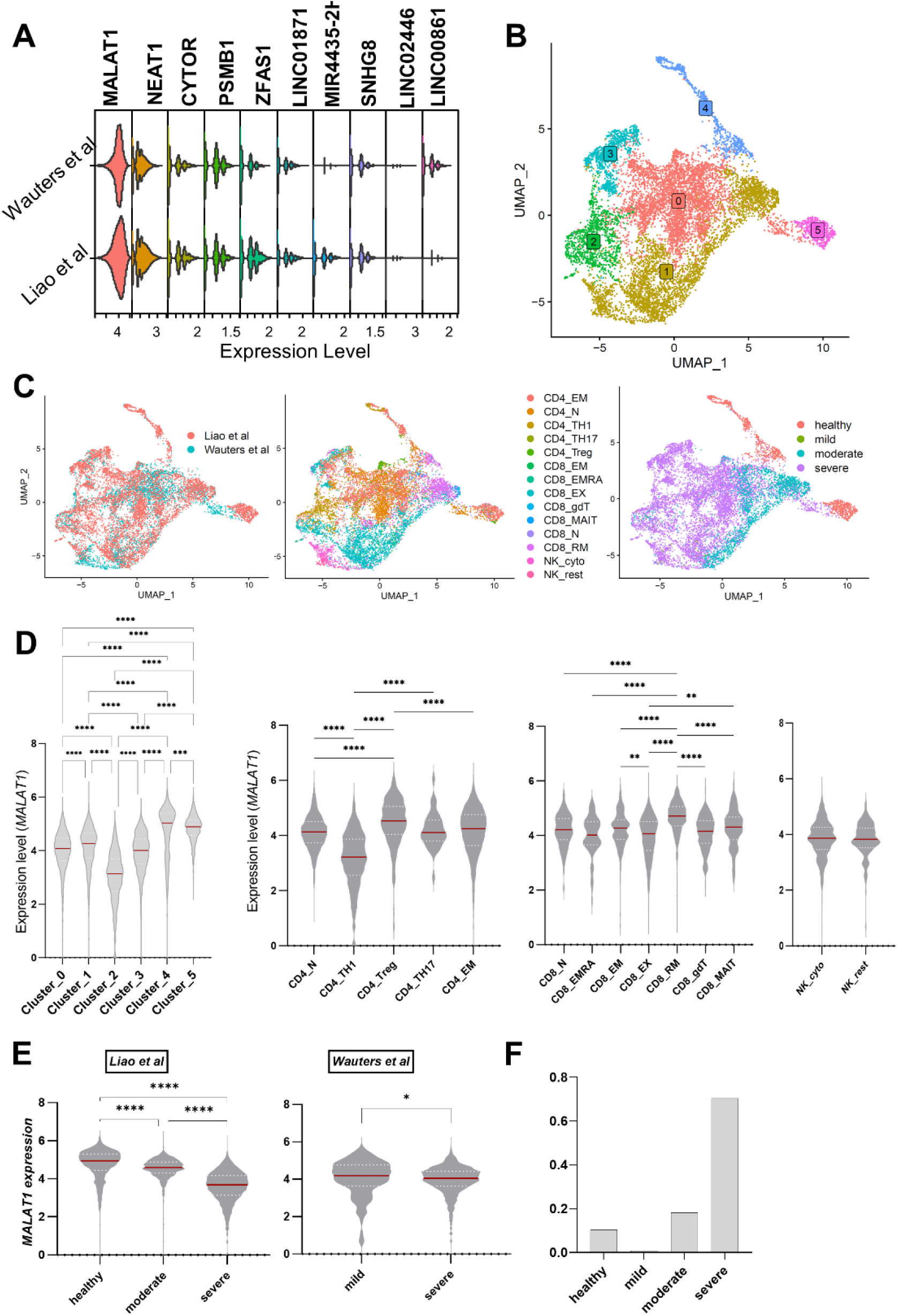
*MALAT1* is differentially expressed in T cell subsets: **A)** Stacked violin plots showing the top 10 highly expressed LncRNA in T cells in bronchoalveolar lavage fluid studies. **B)** UMAP plots depicting single cells coloured by their cluster identity. **C)** UMAP plots depicting single cells coloured and labelled by their respective study (Liao et al., 2020 and Wauters, Mol, et al., 2021). Same as left but coloured by imputed cell sub-population type and disease severity (healthy=*1225*, mild=*111*, moderate= *2135*, severe=*8275* cells) respectively. **D)** Violin plots showing normalised counts of *MALAT1* expression across cluster identities of T cells and imputed cell sub-populations. Kruskal-Wallis multiple comparison p-values are indicated by asterisks (****p<0.0001, ***p=0.0001-0.005,**p=0.005-0.001,*p=0.01-0.05). For cell sub-populations, only a subset of comparisons is shown. **E)** Violin plots showing normalised counts of *MALAT1* expression across study and disease severity. Kruskal-Wallis multiple comparisons with p-value asterisks as defined in **D**. **F)** Barplot showing proportion of total cells represented by each severity group pooled for both datasets.

Importantly, we found that *MALAT1* is differentially expressed within unbiased clusters, especially cluster 2 (**Figure 1D left**) and within imputed T cell sub-populations. We found that Th1 cells (CD4_TH1) demonstrate lower *MALAT1* levels with respect to naïve CD4+ T cells (CD4_N), confirming previous findings in mouse Th cells (Hewitson et al., 2020). CD4_Treg showed the highest *MALAT1* levels. We also observed differences in *MALAT1* expression within CD8+ T cells, with CD8_RM subset showing the highest *MALAT1* expression compared against all other subsets. The difference in *MALAT1* expression between CD8_RM and CD8_EM/CD8_EMRA may mark how a memory T cell be poised towards tissue homing (Kok et al., 2021). While exhausted CD8+ T cells (CD8_EX) had a lower median value of MALAT1 than naïve CD8+ cells (CD8_N), this was not significant. However, compared to CD8_EM, CD8_EX had lower MALAT1 levels. It is notable that the lower quartile of CD8_EX cells was the lowest among all CD8 subsets (**Figure 1D right**). In our data integration (see **Methods**), we retained cell cycle genes, as *MALAT1* has been previously linked to cell cycle (Tripathi et al., 2013; Wang et al., 2016). In doing so, and as suggested (Liao et al., 2020; Wauters, Van Mol, et al., 2021), we found cluster 2 (**Figure 1B and C middle**) to be a mix of CD4_TH1 and CD8_EX T cells (**Figure 1D)**. Interestingly, *MALAT1* expression was reduced in T cells from BAL from severe patients in both the datasets (**Figure 1E**), although we note that this may be biased due to the low proportion of cells from non-severe patients (**Figure 1F**). Interestingly, among the top 10 highly expressed lncRNAs (**Figure 1A, Figure S2**) only *MALAT1* seemed to be down-regulated in severe cases with respect to both healthy, mild/moderate cells (**Figure 1E** versus **Figure S2**).

### MALAT1(anti-)correlated gene lists identify CD8+ T_EX_ CD4+ T_TH1_ cells

To understand the effect of variability in *MALAT1* expression (**Figure 1D**) across coarse- and fine-grained T cell heterogeneity we looked at how *MALAT1* gene expression correlated against all other genes across all T cells, or only CD4+ T cells or CD8+ T cells, respectively. Keeping a significance score of p=0.05 and positive correlation value > 0.1 or negative correlation value < -0.1 as a cut-off, we found that ∼80% of the genes that significantly co-vary with *MALAT1* are those anti-correlated to its expression (all T cells, **Figure 2A**). This percentage is ∼88% for CD4+ T cells and ∼65% for CD8+ cells when correlations were calculated separately for CD4+ and CD8+ cells (**Figure 2A**).

**Figure 2:**
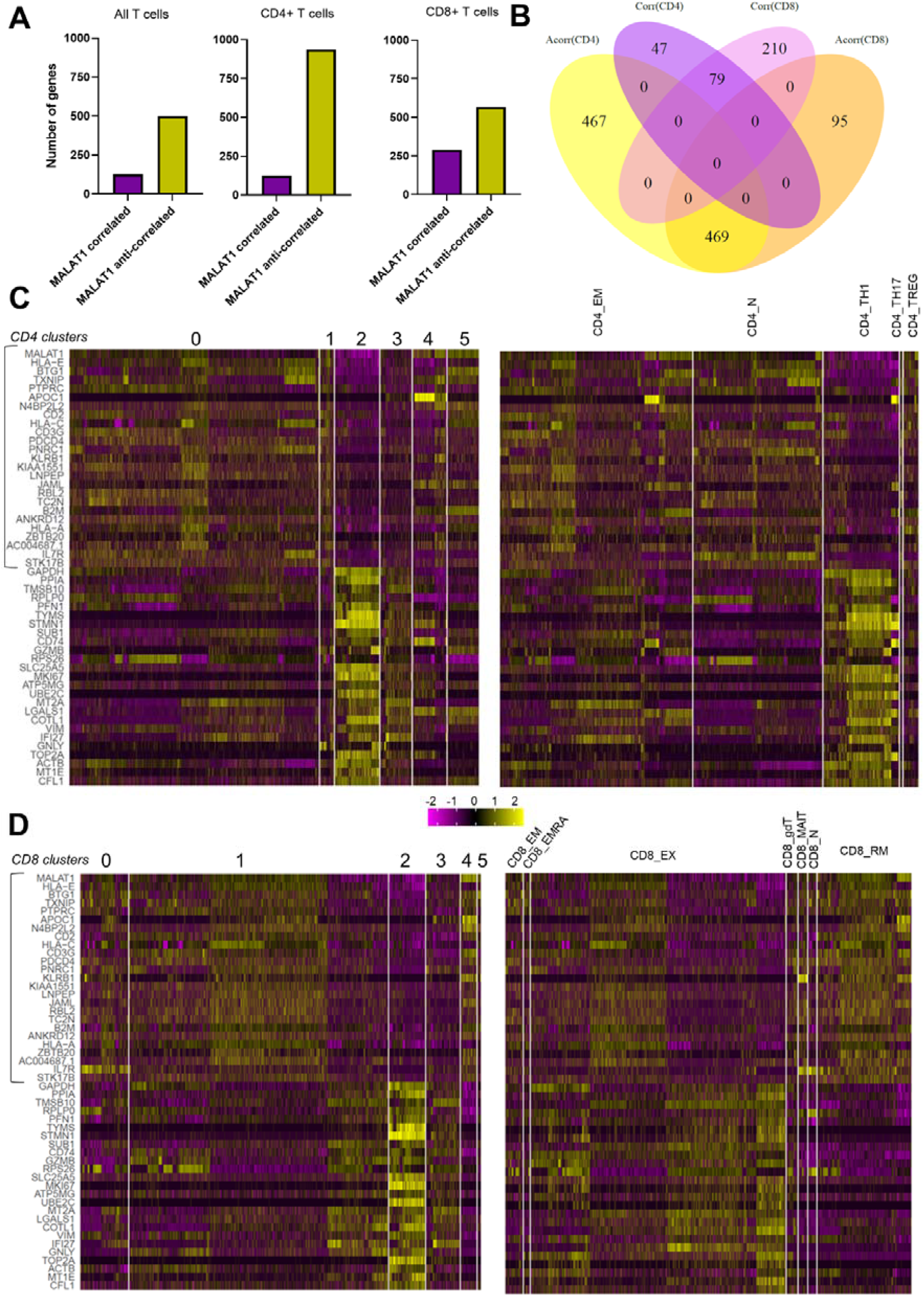
BAL T cell sub-populations differentially express *MALAT1*-correlated genes. **A)** Bar plots showing the number of genes that correlate or anti-correlate with *MALAT1* for all T cells, CD4+ and CD8+ T cells. **B)** Venn diagram depicting the intersection of gene lists that either correlated [Corr(CD8) in violet, Corr(CD4) in purple] or anti-correlated [Acorr(CD8) in light brown, Acorr(CD4) in yellow] with *MALAT1* expression in CD4+ cells OR CD8+ cells. **C)** Heatmap of top 25 *MALAT1* correlating genes (highlighted in the rectangle) and top 25 anti-correlating genes in CD4+ T cells grouped by their cluster identities and by sub-population **D)** Same as C but for CD8+ T cells.

Upon analysing the intersection of these gene lists we found that out of the genes that correlated positively with *MALAT1* for CD4+ and CD8+ cells, there were 79 genes that were common between the two T cell types while there were over four times as many uniquely *MALAT1* correlated genes in CD8+ over CD4+ cells (210 versus 47; **Figure 2B**). While CD4+ and CD8+ cells shared a high number of genes that were anti-correlated to *MALAT1*, the number of genes that were unique in anti-correlation lists were four times as many in CD4+ T cells than CD8+ T cells (**Figure 2B**).

We next identified whether the top 25 *MALAT1*-correlated and top 25 *MALAT1*-anti-correlated genes in both CD4+ T cells (**Figure 2C**) and CD8+ T cells (**Figure 2D**) were differentially expressed in clusters identified previously (**Figure 1D left**) or within imputed cell sub-populations (**Figure 1D centre**). When grouped by cluster identities, in CD4+ T cells, *MALAT1* anti-correlated genes are upregulated in cluster 2 (bottom 25 genes, **Figure 2C**) while a strong *MALAT1* correlated signature is observed in clusters 0, 1 and 5 (top 25 genes, **Figure 2C left**). Cells in cluster 2 (**Figure 2C left**) may predominantly be CD4_TH1 cells (**Figure 2C right**) while cluster 0, 1 and 5 (**Figure 2C left**) may largely comprise of either CD4_N (naïve) or effector memory (CD4_EM) CD4+ T cells (**Figure 2C right**).

In a similar manner in CD8+ T cells, cluster 2 is characterised by genes that are *MALAT1* anti-correlated (bottom 25 genes, **Figure 2D**). When grouped by T cell sub-populations, CD8+ T_EX_ cells appeared to be enriched in the *MALAT1* anti-correlated signature (**Figure 2D**). When grouped by cluster identities, *MALAT1* anti-correlated genes were expressed in cluster 2 of CD8+ T cells (**Figure 2D**) as with CD4+ T cells (**Figure 2C**). Interestingly, CD8+ T_EX_ population appears heterogeneous in terms of expression of MALAT1 anti-correlating and correlating genes (**Figure 2D**) which may explain why MALAT1 expression is not significantly different between CD8 T_N_ and CD8 T_EX_ cells (**Figure 1D right**). In addition, *MALAT1* correlated signature is enriched in resident memory subset (CD8+ T_RM_) and effector memory (CD8+ T_EM_) sub-populations (**Figure 2D**).

Importantly, *MALAT1* anti-correlated with the marker of proliferation *MKI67* in both CD8+ and CD4+ and its expression is increased in cluster 2 (**Figure 2C and D**) potentially indicating the proliferative nature of cells in this cluster. Overall, these findings identified a core gene signature that anti-correlates with MALAT1 expression in T cells and indicated that these genes were highly expressed in proliferative CD4_TH1 and CD8_EX cells.

### MALAT1 anti-correlated genes include a core proliferation and cell specific signature in T cells

Next, we used STRING-DB to perform network analysis for the top 100 genes that anti-correlate with *MALAT1* in both CD4+ and CD8+ T cells as networks (**Figure 3A**). Upon clustering these using k-means (k=3), the resulting clusters showed FDR corrected enrichment for “Cell Division”, “Oxidative phosphorylation” and “Response to Cytokine” (**Figure 3A**).

**Figure 3:**
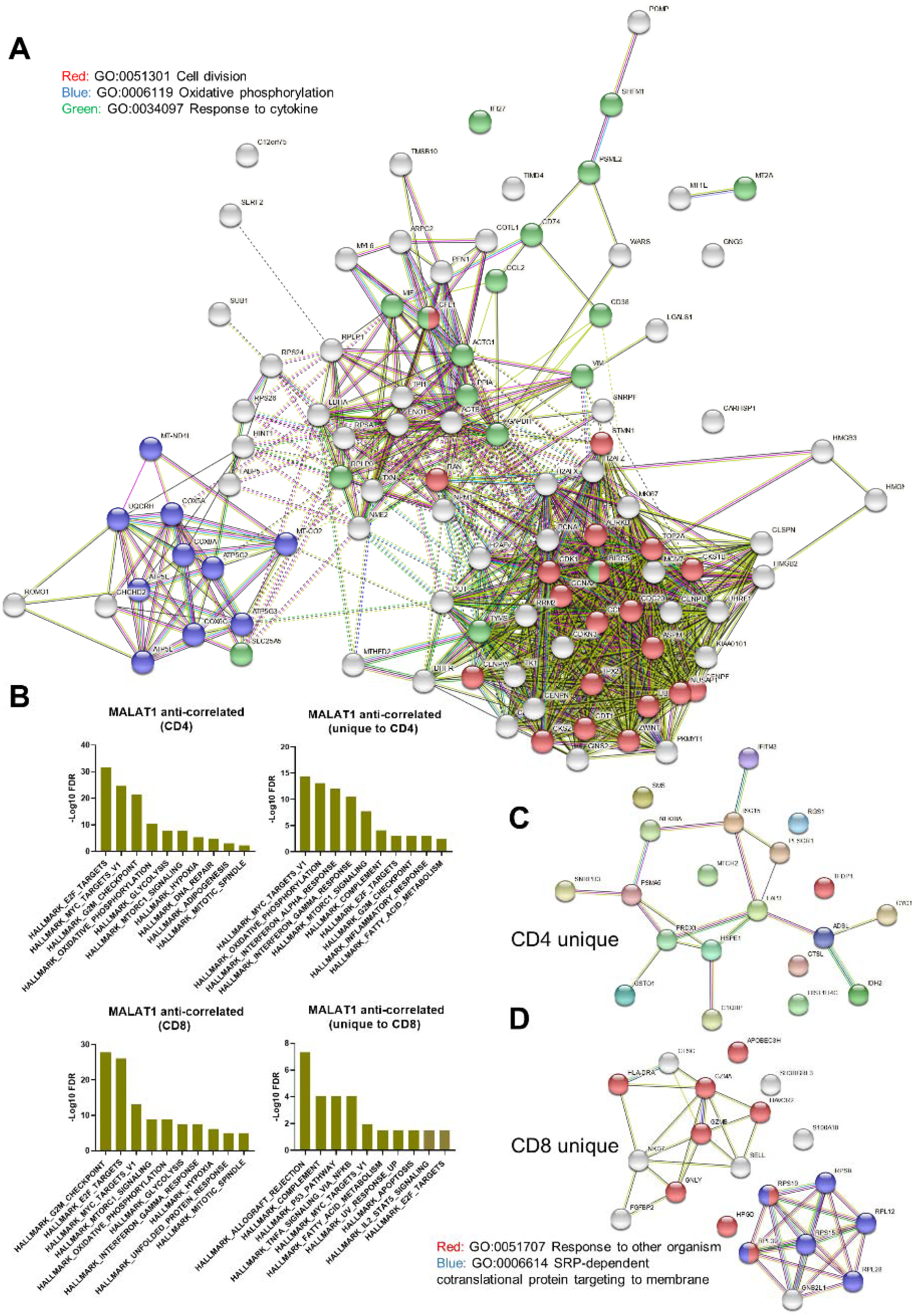
*MALAT1* anti-correlates to genes that are related to cell cycle progression in T cells. **A)** Network representation of STRING interactions plotted for top 100 *MALAT1* anti-correlated genes and those that are common in CD4+ and CD8+ T cells with lines indicating interconnectedness in terms of co-expression or interaction. **B)** Barplots showing negative log FDR values for HALLMARK GSEA enrichment for genes that anti-correlate with MALAT1 and those that are uniquely so in CD4+ and CD8+ T cells. **C)** Same as A but for top 20 unique genes for CD4+ T cells. **D)** Same as D but for CD8+ T cells.

Next, we investigated the gene lists using Gene Set Enrichment Analysis (Subramanian et al., 2005) using the hallmark gene sets to look at signatures within our gene lists. *MALAT1* anti-correlated genes were significantly enriched for HALLMARK_E2F_TARGETS (cell cycle targets of E2F transcription factors), HALLMARK_MYC_TARGETS_V1 (genes regulated by MYC), HALLMARK_G2M_CHECKPOINT (progression through cell division) for CD4+ and CD8+ T cells suggesting the *MALAT1* anti-correlated signature might play a role in proliferation, cell cycle progression and protein translation (CD4 and CD8, Figure 3B). Further, these gene sets showed enrichment for HALLMARK_HYPOXIA, HALLMARK_OXIDATIVE_PHOSPHORYLATION and HALLMARK_GLYCOLYSIS for both CD4+ and CD8+ T cell whereas genes for HALLMARK_DNA_REPAIR only enriched in *MALAT1* anti-correlated gene list for CD4+ T cells (**CD4, Figure 3B**).

HALLMARK_INTERFERON_GAMMA_RESPONSE and HALLMARK_INTERFERON_ALPHA_RESPONSE (genes upregulated in response to IFN-γ and IFN-α signalling) were hallmarks uniquely associated with CD4+ T cells (**unique to CD4, Figure 3B**). HALLMARK_COMPLEMENT is associated with both CD4+ and CD8+ T cells with HALLMARK_XENOBIOTIC_METABOLISM associated with CD4+ T cells (**unique to CD4, Figure 3B**). Further, *MALAT1* anti-correlating genes uniquely in CD8+ T cells were enriched for genes involved in the p53 pathway (HALLMARK_P53_PATHWAY) and those regulated in response to TNF via NF-κB (HALLMARK_TNFA_SIGNALING_VIA_NFKB, **unique to CD8, Figure 3B**).

We looked at the top 20 genes uniquely anti-correlated to *MALAT1* in CD4+ T cells for STRINGDB interactions and found that genes related to response to TNF/IL-1 such as PSMA5, NFKBIA and Ubiquitin cross-reactive protein (ISG15) (**Figure 3C**). On the other hand, in CD8+ T cells, MALAT1 uniquely anti-correlates with genes associated with membrane targeting of proteins along with genes involved in CD8+ T cell exhaustion like GNLY, GZMB, GNLY and HAVCR2 (**Figure 3D**).

### MALAT1 and MKI67 anti-correlate in COVID-19 post-mortem lung tissue

The above cell-type gene signature and pathway analyses indicated a potential link between *MALAT1* expression and T cell proliferation. To further test this, we checked if the *MALAT1* anti-correlated gene list signature common to CD4+ and CD8+ T cells (**Figure 3A**) identified in BAL samples was sufficient to mark proliferating T cells in lung tissue. For this purpose, we analysed a COVID-19 explant/post-mortem lung scRNA seq dataset (Bharat et al., 2020). We pre-filtered barcodes labelled as “T cells” from the dataset and used the top 100 common genes that anti-correlated with *MALAT1* (**Figure 3A**), of which 88 genes were found in Bharat et al, to calculate the ‘area under recovery curve’ or AUC (Aibar et al., 2017) for each cell (**histogram, Figure 4A**) to identify gene list enrichment. Thresholding the AUC score (at AUC>=0.39) based on the bimodality in AUC distribution (**histogram, Figure 4A**), the cells were highlighted on a UMAP plot (**Figure 4A**). The high AUC score highlights proliferating T cells as indicated by their corresponding *MKI67* expression and lower *MALAT1* levels (**Figure 4B**).

**Figure 4:**
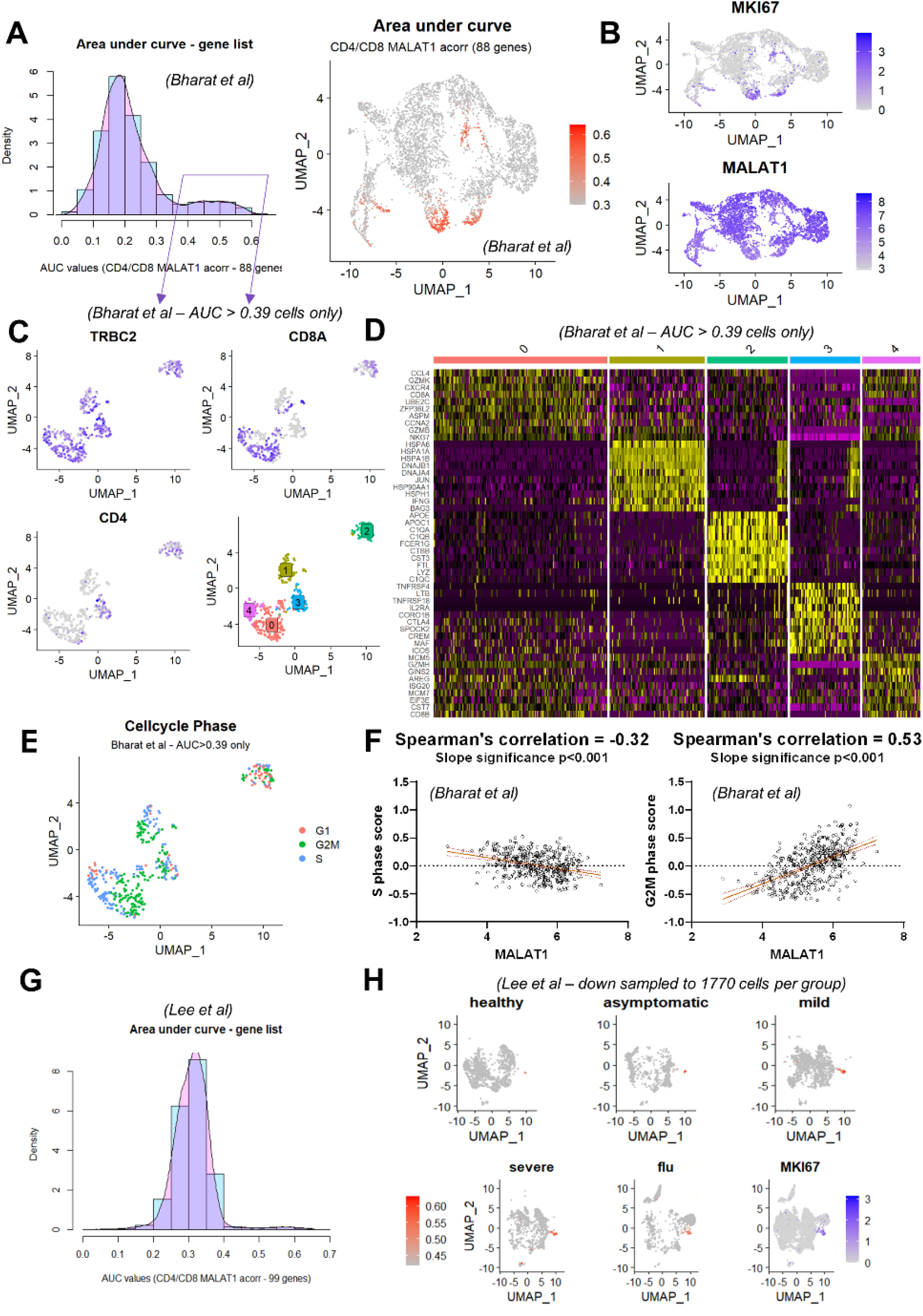
*MALAT1* expression is reduced in proliferating T cells from lung digests. **A)** Histogram showing ‘area under recovery curve’ score for *MALAT1* anti-correlated gene list (number of genes=88) for single cells. UMAP plot highlighting cells with AUC score from **B** that are greater than 0.39 **B)** UMAP plot depicting MKI67 and *MALAT1* expression per single cell. **C)** UMAP plot depicting TRBC2, CD8A and CD4 gene expression along with cluster identities of proliferative T cells (subset based on AUC score greater than 0.39 from **A**). **D)** Heatmap showing top 10 genes expressed in each imputed cluster in **C** **E)** UMAP plot depicting imputed cell cycle phase for each T cell. **F)** Scatter plots and linear regression between imputed S-phase score (left) and G2/M score (right) per cell and *MALAT1*. P-values indicates the significance of the slope of the regression. **G)** Histogram showing ‘area under recovery curve’ score for *MALAT1* anti-correlated gene list (number of genes=99) for PBMCs. **H)** UMAP plot highlighting cells with AUC score in PBMCs from different groups along with MKI67 expression where each dot represents a single cell.

Interestingly, the proliferative T cells in post-mortem lung tissue appeared diverse in terms of their position in UMAP space. To investigate this further, we examined a subset of these cells (AUC scores >= 0.39) and visualised canonical T cell markers in two-dimensional UMAP space (**Figure 4C**). We further re-clustered these cells (**Figure 4C**, bottom right) to understand whether the heterogeneity in proliferative T cells with a high AUC score (**Figure 4A**) translates in terms of differential gene expression (**Figure 4D**).

We found that that the largest proliferative cluster (cluster 0, **Figure 4C**) comprised of CCL4, CXCR4 expressing CD8+ T_EM_ (CD8A, GZMK, GZM, MKG7, GZMH) cells (**Figure 4D, Supplement ST1**). Cluster 1 was comprised of IFNG+ gdT cells (**Figure 4D**) expressing TRDC and GNLY (**Supplement ST1**). Cluster 2 markers appeared to have a strong macrophage-like gene signature with complement genes, FCER1G, LYZ being upregulated in this cluster (**Figure 4D**). As this cluster also expressed T cell markers (**Figure 4C**), we postulated that these were doublets and were not further analysed. Cluster 3 was enriched in markers for CD4 T_REG_ such as IL12RA and CTLA4 (**Figure 4C**) whereas cluster 4 comprised of CD8+ (CD8B, **Figure 4C**) cells with targets of E2F transcription factors such as MCM5.

Since our analysis suggested a decrease in *MALAT1* was linked to a corresponding increase in *MALAT1* anti-correlating genes, we asked if *MALAT1* expression levels were consistently lower in proliferative cells and if this was dependent on their cell cycle state. We calculated a score based on genes involved in cell cycle progression (Nestorowa et al., 2016) including those involved in the S, G2/M and G1 phases. In **Figure 4E**, we show how these proliferative T cell clusters (**Figure 4C**, bottom right) comprise of cells in S, G2/M and G1 phase. We then calculated Spearman’s correlation between imputed cell phase score and *MALAT1* and found that *MALAT1* levels anti-correlated with the imputed S phase score and strongly positively correlated with the G2M score (**Figure 4F**).

To test if our findings were limited to tissue T cells, we examined a COVID-19 PBMC dataset (Lee et al., 2020). Using the above-defined top 100 genes that anti-correlate with *MALAT1*. We identified a small proportion of cells within this dataset that expressed these genes differentially (99/100 genes were found in the dataset, **Figure 4G**). In fact, this signature also picks out PBMCs from influenza patients, suggesting that this is a hallmark feature of T cells responding to infection. As in the case of lung T cells, these AUC > 0.42 cells (**Figure 4G,H**) are found to be neighbourly in UMAP space and express MKI67 (**Figure 4H**).

To test the above findings *in situ*, we examined post-mortem lung sections from the UK Coronavirus Immunology Consortium (UK-CIC) (patient_meta_data, **Supplement ST1 and Milross et al, ms in prep**). We analysed lung autopsy sections (n=6) and representative sections stained with DAPI are shown in **Figure 5A**. We then determined CD8 expression and MKI67 (as a marker for proliferation) by immunofluorescence along with *MALAT1* by RNAScope. We found, qualitatively, that MALAT1 was seldom co-expressed with MKI67 unless MKI67 levels were high (**Figure 5B, C & D**). Interestingly, this suggested some correlation of MALAT1 and MKI67 when the latter was more highly expressed. This may be related to the *MALAT1* expression correlation we observed with the G2M phase T cell score (**Figure 4F**). We next performed quantitative analysis using QuPath (**Figure 5E**). For all tested samples we observed distinct MKI67-hi/MALAT1-lo populations, with the majority of highest MKI67 expressing CD8+ T cells (mean nuclear intensity > 2000) showing low *MALAT1* levels. We also found double positive CD8+ cells that co-express *MALAT1* and MKI67 (**Figure 5F**). These double positive cells may be explained by the particular phase of the cell, as it has been shown that MKI67 is not a binary marker for proliferation but a graded marker for proliferation/senescence (Miller et al., 2018). In general, however, we found that when MALAT1 expression is high then MKI67 expression is low and vice versa (**Figure 5F**) across all tested samples.

**Figure 5:**
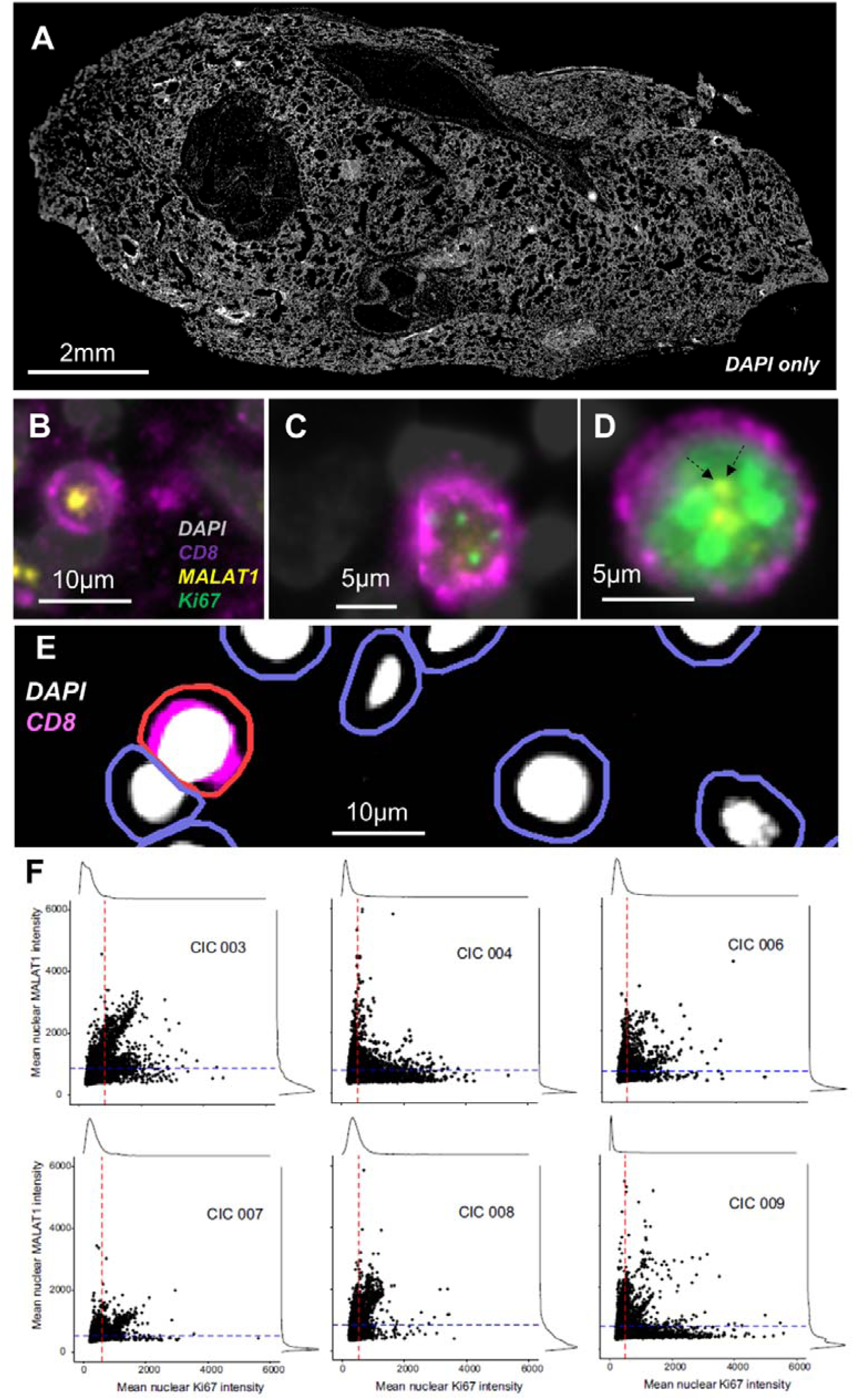
*MALAT1* expression is reduced in proliferating CD8+ T cells post-mortem tissue. **A**. DAPI stained (grey) whole lung autopsy section. **B-D**, Immunofluorescence and RNAScope images showing nucleus in grey (DAPI), MKI67 protein in green, CD8 surface protein in purple and *MALAT1* RNA in yellow. Images show representative cells negative for MKI67 and positive for MALAT1 (B), weakly positive for MIK67 and with undetectable MALAT1 (C) and highly expressing MKI67 along with *MALAT1* (D). Dotted arrows in (D) highlight MALAT1 positive signal for clarity. **E**. Strategy employed to detect cells wherein first nucleus was identified and then cell boundaries using QuPath (see *Methods*) and a positive cell was detected (shown here in red) based on CD8 fluorescence intensity of the cell. **F**. Scatter/histogram plots per autopsy along with horizontal and vertical lines drawn to the 3^rd^ quantile of mean nuclear MALAT1 and MKI67 intensity respectively matched to the histogram. The two straight lines divides the plot into four quadrants of relative expression (see *Methods*).

## Discussion

*MALAT1* is one of the most abundant non-ribosomal RNA transcripts in mammalian transcriptomes. Despite an increasing understanding of how *MALAT1* upregulation contributes to cancer development and progression (Goyal et al., 2021), less is known about its physiological functions in non-transformed cells. Recent work from our and other laboratories has indicated that MALAT1 plays a role in T cell function and that, in preclinical models, antigenic activation of naïve T cells results suppression of *MALAT1* expression (Hewitson et al., 2020; Kanbar et al., 2022; Liang & Tang, 2020; Masoumi et al., 2019; Xue et al., 2022). Here, we used published annotated transcriptomic datasets in COVID-19 to specifically look at T cell phenotypes ranging from naïve to effector memory and exhausted and found that *MALAT1* is negatively correlated with a core gene signature in T cells, which in turn is linked to cellular proliferation. Using post-mortem lung autopsy samples, we experimentally validated this association and showed that MKI67+ proliferative CD8+ cells are characterised by low *MALAT1* expression.

T cell proliferation can be spontaneous or homeostatic (Min et al., 2005) and the conditions that regulate the same varies between CD4+ and CD8+ T cells (Do & Min, 2009). CD8+ T cell proliferation is essential with rapid proliferation in response to interaction with a foreign peptide but also during homeostasis if T cell numbers fall below a threshold (Min, 2018). The former, however, progresses through to a CD8 effector memory phenotype (Surh & Sprent, 2000). In fact, it has been demonstrated in CD8+ T cells, that a T central memory phenotype is marked by a higher number of prior divisions than the effector memory T cell pool (Bresser et al., 2022; Obar & Lefrançois, 2010; Sarkar et al., 2008). The replicative history of T cells is closely connected to its functional repertoire (Bresser et al., 2022). Interestingly, CD8+ T exhausted cells in COVID-19 are connected via the CD8 T_N_, CD8+ T_EM_ lineage (using pseudotime analysis) and have higher levels of proliferation markers (Wauters, Mol, et al., 2021). Indeed, using Wauters, Mol, et al., 2021 dataset, we note that CD8+ T_EX_ have a corresponding lower *MALAT1* level with increased expression of *MALAT1* anti-correlating genes (**Figure 2C**).

*MALAT1* has been long associated with enhanced proliferation in cancer (Gutschner et al., 2013; Li et al., 2009) and the lack of the gene is shown in human diploid lung fibroblasts to have a reduction in their proliferation with an arrest at the G1/S phase with an increase in genes involved in the p53 pathway (Tripathi et al., 2013). Interestingly, in T cells we observe a physiological downregulation of *MALAT1* that anticorrelates with the S phase score of cells (**Figure 4F**), suggesting *MALAT1* suppresion may be a consequence of T cell proliferation. Interestingly, overall *MALAT1* levels anti-corelate to HALLMARK_P53_PATHWAY (**Figure 3B**) and is unique to CD8+ T cells. While *MALAT1* in this work has been shown to anti-correlate with a cell’s S phase score (**Figure 4F**), it has been shown that many lincRNAs peak during S phase in human epithelial cells leading to transcriptional regulation during cell cycle progression (Yildirim et al., 2020). In that study it was found that *MALAT1* peaks at close to the beginning of G2/M (Yildirim et al., 2020). In this respect, we found *MALAT1* levels to correlate with G2M score in T cells (**Figure 4F**), which indicate similarity of T cells to epithelial cells in terms of *MALAT1* expression during cell cycle.

We find that more genes anti-correlate with *MALAT1* than those that correlate (**Figure 2A**). Whether this is due to direct effects of *MALAT1* through its roles in gene regulation (Arun et al., 2020) will need to further tested. However, it suggests that physiological regulation of *MALAT1* levels may alter gene expression of T cells that are known for their plasticity (Dupage & Bluestone, 2016). Further still, as cell proliferation is central to T cell activation (Hwang et al., 2006; Yoon et al., 2010), it will be interesting to investigate how a lack of *MALAT1* during proliferation may shape T cell function upon subsequent activation and differentiation. We have previously reported that lack of *MALAT1* results in lower levels of MAF and IL10 in mice and as a consequence, greater host resistance to infection or increased immunopathology (Hewitson et al., 2020). Others have reported impaired CD8+ T cell function upon MALAT1 loss (Kanbar et al., 2022). Interestingly, MALAT1 mediates its function through interactions with proteins and potentially RNA, interactions which based on the results presented here would be expected to be altered in MALAT1-lo proliferating T cells. Genes that may be associated with shaping T cell function post proliferation may indeed lie amid the *MALAT1* anti-correlated signature that we find in CD4+ and CD8+ T cells, especially those involved in cytokine response and oxidative phosphorylation (**Figure 3A**). As an example we find HAVCR2 (TIM-3) which is a marker for T cell exhaustion (Tang et al., 2019) anti-correlates with *MALAT1* (unique to CD8+ T cells, **Figure 3D**). How these genes may vary between T cell subsets such as CD8+ T central memory where lowly divided cells are capable of mounting a better effector response upon re-infection (Bresser et al., 2022) and exhausted CD8+ T cell population with increased cell cycle markers like MKI67 (Z. Chen & Wherry, 2020; Wauters, Mol, et al., 2021) requires further investigation.

Taken together our results reveal that suppression of *MALAT1* expression is a feature of proliferating activated T cells. This means that *MALAT1*-associated functions are likely to be suppressed in proliferating T cells, but not necessarily that MALAT1 suppression drives the proliferation. The *MALAT1*-linked gene signatures identified here provide an initial insight into the potential functional consequences of *MALAT1* suppression in human T cells, forming the foundation for further mechanistic studies on the function of this highly expressed lincRNA in T cells within and beyond viral infection.

## Methods

### Datasets

Single-cell RNA seq data from healthy and COVID-19 patients from Gene expression omnibus accession number GSE145926 and is referred throughout the paper as Liao et al (Liao et al., 2020) and T cell barcodes (using metadata from original publication) were subset and used further for analysis. Dataset Wauters et al (Wauters, Van Mol, et al., 2021) was obtained from https://lambrechtslab.sites.vib.be/en/data-access. Specifically, the file T_NKT_cells.counts.rds was downloaded to use as counts matrix. Single cell data for barcodes with ‘COVID19’ as metadata were included in the downstream analysis from the Wauters et al dataset.

Finally, accession number GSE158127 (Bharat et al., 2020) was used to analyse post-mortem T cells form lungs and for validation. Further, GSE149689 (Lee et al., 2020) was used to look at *MALAT1* signatures in PBMCs and in flu.

### In silico T cell quality check and phenotype identification

Single T cell transcriptomes from Liao et al and Wauters et al were loaded as Seurat (v4.0.5) objects and the latter Seurat object’s metadata describing T cell phenotypes were used to impute T cell phenotypes in Liao et al using the functions FindTransferAnchors() and Transferdata(). Next, single transcriptomes with greater than 5% mitochondrial genes were discarded from downstream analysis. Next, counts from both Seurat objects were regressed using percentage of mitochondrial genes, ribosomal genes, total RNA count and number of unique features using method “glmGamPoi” which is available as an R package with the same name (https://bioconductor.org/packages/release/bioc/html/glmGamPoi.html). Finally, the anchors between the two Seurat Objects were found (functions SelectIntegrationFeatures() and FindIntegrationAnchors()) to then integrate (IntegrateData()) them into a single integrated Seurat object.

### Dimensionality Reduction

Principal components analysis was performed on the integrated Seurat object (3000 variable features). Top 30 PCA components were used to cluster the data by a K-nearest neighbour clustering using FindClusters() with a resolution parameter of 0.8. UMAP was performed on the PCA space and single cells were represented on UMAP axes and coloured by their cluster membership.

### Correlation analysis

Correlation of all genes with *MALAT1* was calculated using cor.test() from the stats package in R (4.1.1) implemented with Spearman’s ranked correlation method. The level of significance associated with a correlation was set at 0.05. Correlation values between -0.1 and 0.1 (both included) were excluded.

### Network & Gene set enrichment analysis

Network analysis of gene lists were performed on String-DBGene set enrichment analysis was performed on STRING (https://string-db.org/). Gene set enrichment analysis (GSEA, http://www.gsea-msigdb.org/gsea/msigdb/annotate.jsp) using the option to ‘Investigate Gene Sets’ to search for significant (*p-value corrected*) overlaps with Hallmark gene sets, GO biological process, cellular component and molecular function.

### Area under curve

Gene list enrichment in cells was calculated using the R package AUCell, originally published as a part of SCENIC (Aibar et al., 2017). Expression matrices as obtained from the Seurat object were provided to the function AUCell_buildRankings() to build cell rankings which was then was used to calculate a ‘area-under-recovery-curve’ for the provided gene list. AUC score thresholds were selected based on visual inspection and are indicated in the relevant figure.

### RNAScope and immunofluorescence

Post-mortem autopsy sections from UK-CIC first wave cohort (CIC003-9) were obtained on glass slides and stained for CD8, MKI67 and DAPI. *MALAT1* was probed on the same section using RNAScope (Bio-techne) FISH assay as per manufacturer’s instructions.

### QuPath

All images were acquired on a Zeiss AxioScan.Z1 slide scanner. Exposure times and threshold settings for all three channels were used for each of the images. Images in the CZI format were loaded on QuPath-0.3.2 (Bankhead et al., 2017). Whole images were analysed for co-expression of MKI67, CD8 and *MALAT1* at single-cell resolution and count data was analysed. CD8+ cells were detected using the module ‘positive cell detection’ using DAPI as a counterstain to draw nuclei and cell boundaries. Cellular CD8 intensity was then used to detect positive cell types. Data was exported and then further investigated in R. Cells with a circularity score of less than 0.75 were excluded and expression positivity for MKI67 and MALAT1 was determined by selecting only those cells that had a maximum pixel intensity greater than the minimum detected intensity.

### Study/ethics approval

Human lung tissue samples from individuals who died of COVID-19 during the first wave of the pandemic in the UK were obtained from the Newcastle Hospitals CEPA Biobank and their use in research was covered by Newcastle Hospitals CEPA Biobank ethics – REC 17/NE/0070. Ethics approval was granted by the NHS Research Ethics Service.

### Data availability

All single cell RNA seq data used in this study is publicly available and their source is described in methods under the sub-section “Datasets”. Code used to analyse data is available here - https://github.com/jipsi/malat_proliferation/ and all processed/unprocessed Rds files used in this study are available at - https://doi.org/10.5281/zenodo.7506637. Images and analyses generated by this study can be made available upon request.

## Data Availability

All single cell RNA seq data used in this study is publicly available and their source is described in methods under the sub-section 'Datasets'. Code used to analyse data is available here - https://github.com/jipsi/malat_proliferation/ and all processed/unprocessed Rds files used in this study are available at - https://doi.org/10.5281/zenodo.7506637. Images and analyses generated by this study can be made available upon request.

https://doi.org/10.5281/zenodo.7506637

## Author Contributions

SD performed formal investigation and analyses. HA performed RNAscope experiments. DL, SD, and PMK, conceived the study. AJFilby, AJFisher, LM, BH, JM performed post-mortem sample collection, tissue handling, and diagnoses. SD and DL wrote the manuscript. All authors edited the manuscript.

## Conflict of interest disclosure

The authors declare no conflicts of interest.

## Funding

This work was funded by UK Research and Innovations / NIHR UK Coronavirus Immunology Consortium (UK-CIC; MR/V028448). PMK is also supported by a Wellcome Trust Senior Investigator Award (WT104726).

## Acknowledgements

We are truly grateful to the families of the individuals who died of COVID-19 for consenting for provision of post-mortem samples. We thank staff at the Imaging and Cytometry Lab in the University of York Bioscience Technology Facility for technical support and advice. Graphical summary was created by www.biorender.com.

## Notes

### Competing Interest Statement

The authors have declared no competing interest.

### Author Declarations

Human lung tissue samples from individuals who died of COVID-19 during the first wave of the pandemic in the UK were obtained from the Newcastle Hospitals CEPA Biobank and their use in research was covered by Newcastle Hospitals CEPA Biobank ethics REC 17/NE/0070. Ethics approval was granted by the NHS Research Ethics Service.

