## Supplementary Figures 1 and 2 for "Down-regulation of *MALAT1* is a hallmark of tissue and peripheral proliferative T cells in COVID-19"

Short running title: Low MALAT1 is a hallmark proliferative T cells in COVID-19

Keywords: T cell, proliferation, MALAT1, COVID-19, lncRNA

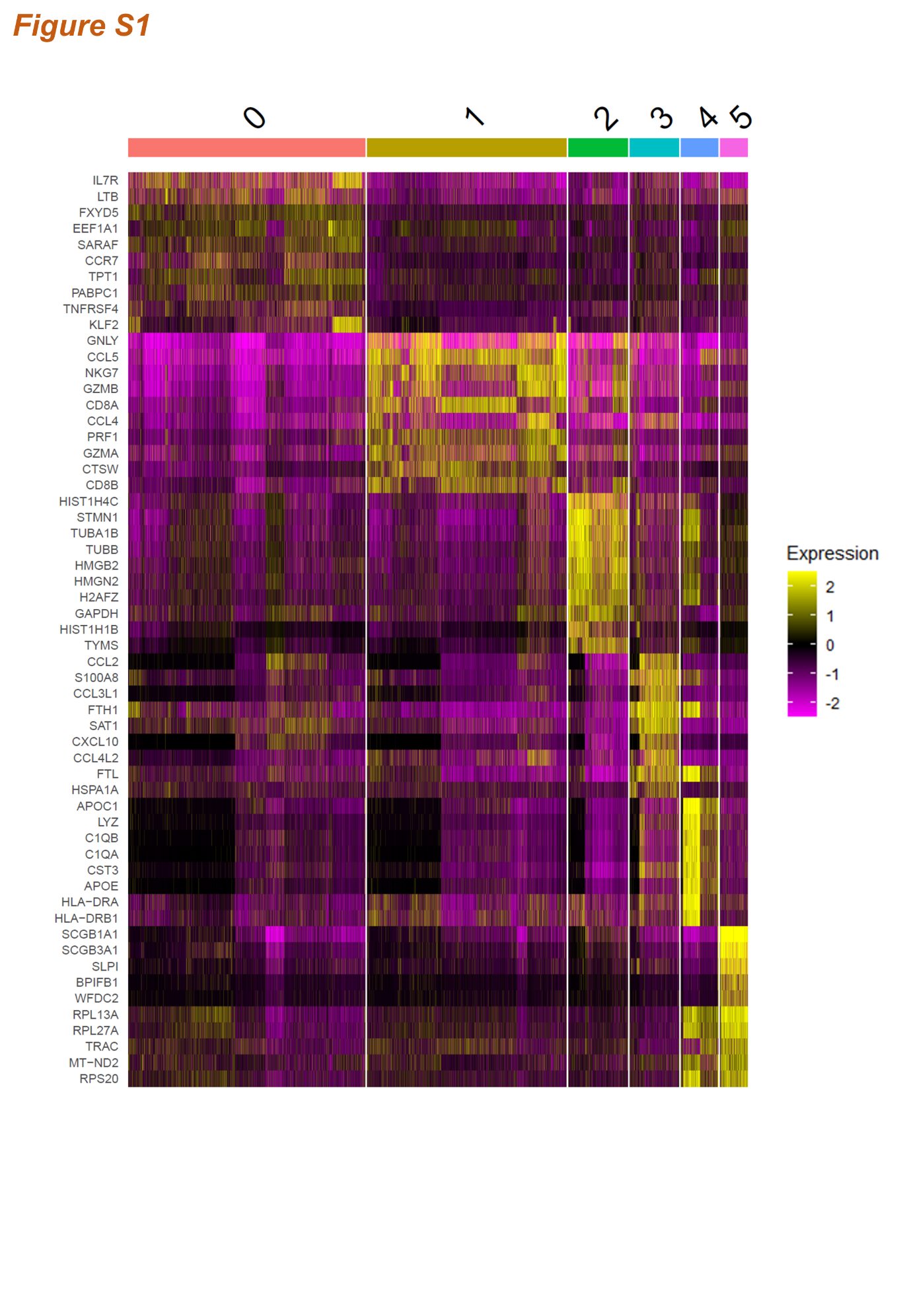

**Figure S1: Gene expression across imputed cell clusters**

Heatmap showing top 10 genes expressed in each cluster as identified in the integrated dataset.

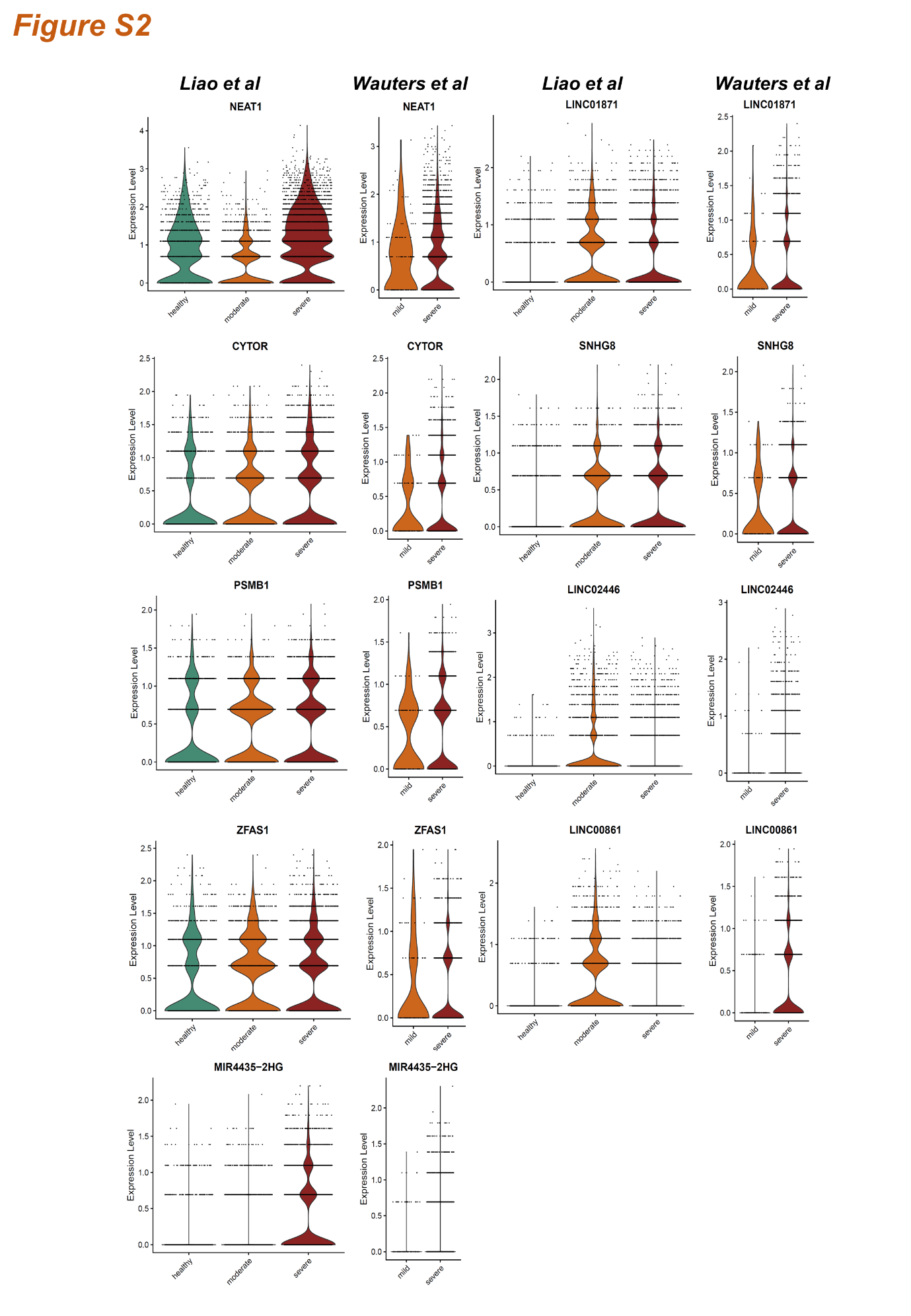

**Figure S2: LncRNA expression across disease severity and dataset**

Violin plots indicating gene expression per cell for the top 2-10 (excluding *MALAT1*) highly expressed lncRNAs in the integrated dataset.
